# PAIN RELIEF IN MAJOR AMPUTATION (PRIMA) PROTOCOL: A SINGLE CENTRE RCT COMPARING PRE-INCISION SINGLE-SHOT NERVE BLOCK AND PATIENT CONTROLLED ANALGESIA TO PRE-INCISION PERI-NEURAL CATHETER, IN PATIENTS UNDERGOING MAJOR LOWER LIMB AMPUTATION

**DOI:** 10.1101/2024.11.21.24317718

**Authors:** L. Shelmerdine, R. Bentley, I Baxter, S Nandhra

**Author notes:** Corresponding Author: Mrs Lauren Shelmerdine, Vascular Registrar. **Chief Investigator** No financial or competing interests to declare. **Principal Investigators** No financial or competing interests to declare.

## Abstract

**Introduction:** 9663 major lower limb amputations were performed in UK NHS hospitals, between 2018-2020. Despite this high number, there is no universally accepted peri-operative analgesia regime. The Vascular Society and Vascular Anaesthesia Society of Great Britain and Ireland, in partnership with patients (supported by the James-Lind Alliance), have identified improving outcomes (including preventing/treating pain) for patients who undergo amputations as a key research priority.

**Methods and methods:** A prospective, single-blind, RCT (1:1), comparing pre-incision ultrasound sited PNC (7-day duration) or pre-incision ‘single-shot’ nerve block and PCA, for those undergoing MLLA. The sample size is 34 patients, powered to detect a primary outcome of 2.5cm(2cmSD) difference on a visual analogue scale (VAS) at day 3.

Secondary outcomes include daily pain scores, analgesia use, post-operative nausea and vomiting, Pasero opioid-induced sedation scale and physiotherapy progress. Patient-reported neuropathic pain and quality of life tools (SF36 and EQ5D) are recorded at baseline, day-7, 6-weeks and one-year.

**Ethics and dissemination:** This study was approved by South East Scotland Research Ethics Committee on 03/02/2021. REC reference:21/SS/0013). It is hoped this NIHR-portfolio adopted, RCS(Ed) funded RCT, will provide level-1 evidence for a shared patient and clinician research priority. Trial registration: ISRCTN.com, ISRCTN64207537. Registered on 21/07/2021

## 1 Background

Major lower limb amputations (MLLA), either transfemoral / above knee amputation (AKA) or transtibial / below knee amputation (BKA), are commonly performed operations by Vascular Surgeons for patients where there is no other option to save the limb, control pain or prevent the spread of infection. In the UK, 500-1000 patients per million population have clinically significant peripheral arterial disease (PAD), 1-2% of whom eventually require MLLA.(1) 9663 MLLA were performed in the UK between 2018-2020.(2) The numbers of MLLA performed each year are likely to increase, as the rates of diabetes increase.(3)

Perioperative pain control is a challenge in these patients, with no universally accepted analgesia regime.(4, 5) In general terms a multimodal approach is recommended for post-operative pain control (6) including opioid sparing analgesics (paracetamol and NSAIDs), opioid analgesics, Patient Controlled Analgesia systems (PCAs) and other adjuvants such as anti-depressants or anti-epileptics, as well as regional analgesia. These include neuroaxial and / or peripheral nerve blockade. Ultrasound guided regional peripheral techniques take place before the start of the operation or ‘pre-incision’ and include single shot blocks (perineural injection of local anaesthetic) or placement of a perineural catheter (PNC). PNCs can be left in situ, instilling local anaesthetic, for varying lengths of time. The surgeon can also site a PNC along the perineural sheath, under direct vision, intraoperatively.

In spite of the range of technique highlighted, it is known that opioid based analgesics are commonly heavily replied upon.(7) In a report into MLLA care by NCEPOD in 2014, post-operative pain relief was only assessed as being ‘good’ in just over a third (37.5% 174/464) of patients.(1) Opioid analgesics are associated with risks including drowsiness, respiratory depression, nausea, vomiting urinary retention and constipation. Delirium rates are also increased, which may increase a patient s length of stay in hospital.(8) Reducing the usage of morphine-based painkillers, could benefit patients. There is limited evidence that the use of (surgical sited) PNCs can reduce opioid consumption in the post-operative period.(9)

Poorly controlled acute post-amputation pain has a strong correlation with chronic pain conditions (chronic stump pain (CSP) and phantom limb pain (PLP)), with the incidence of pain after amputation estimated to be between 30-80%, up to 20 years following an amputation.(7, 10)

There is clearly an appetite to investigate pain within this setting, from both patients and clinicians. The results of the James-Lind Alliance Priority setting partnership (JLA-PSP) exercise identified ‘*how can we improve outcomes for patients undergoing major lower limb amputation?*’ and ‘improving pain’. (11, 12)

The Society of Vascular Anaesthetists (VASGBI JLA-PSP) conducted a similar process in 2015, revealing that two of the top ten priorities for patients and clinicians are to understand ‘*what can we do to stop patients developing chronic pain after surgery?* ‘and ‘*what outcomes should we use to measure the ‘success’ of anaesthesia and perioperative care?*’.(13)

There is evidence to suggest that siting a PNC and starting peripheral nerve blockade pre-incision leads to the prevention of PLP,(14) yet, there is a paucity of good quality evidence surrounding pre-incision US guided PNC use. There is very little published surrounding their use in MLLA at all; a handful of small studies or abstracts and no RCTs. (14-18)

There is a systematic review and meta-analysis updated in 2021, which focused on surgeon sited PNCs placed during amputation under direct vision when compared to placebo or other analgesic regime, (excluding studies including US guided pre-incision PNCs or single shot nerve blocks). Ten studies were included, totalling 731 patients. 350 had a surgeon sited PNC and 381 had standard care. Outcomes pertaining to postoperative acute pain scores, opiate requirements, in hospital mortality and in the longer term CSP and PLP were included.(9) The authors concluded that surgeon sited PNCs lead to lower acute pain scores in the post amputation period and this effect is maintained during subgroup analysis for the studies which were RCTs, but that further RCTs were required to determine any effect of PNCs on PLP or CSP. Three of the authors of this systematic review were key authors of the PLACEMENT trial’s protocol published in 2017.(19) Their feasibility study compared surgeon sited PNC to usual care (standard anaesthetic (general, epidural, spinal, regional) and postoperative analgesia. Pre-incision regional nerve blocks or PNCs were permitted in either group. This trial proved feasibility and a full RCT is close to commencing.(20)

Furthermore, although a single shot intraoperative perineural injection is found in lists of recommended multimodal peri operative analgesics, good quality studies into their efficacy in MLLA are also lacking. One retrospective study from 2011 set out to investigate four analgesic modalities and their effect on chronic pain. Whilst they didn’t find any effect on either CSP or PLP they concluded that those patients who received a peripheral nerve block (or epidural) perceived significantly less pain in the week after surgery compared with the other two methods (NRS [SD] values, 2.68 [1.0] and 2.70 [1.0], respectively).(21)

The PRIMA study aims to address the JLA-PSP research priority areas by understanding the impact of pre-incision US guided pain control on MLLA outcomes and will directly compare two methods for which good quality evidence is lacking. PRIMA will investigate outcomes including acute post-operative pain, as well as additional analgesia use, patient recovery, quality of life and chronic pain.

This protocol has been developed in line with the Standard Protocol Items, Recommendations for Interventional Trials (SPIRIT) 2013 statement.(22)

## 2 Materials and Methods

### 2.1 Design

A prospective, single-blind, RCT (1:1), comparing pre-incision ultrasound sited PNC (7-day duration) or pre-incision ‘single-shot’ nerve block and PCA, for those undergoing MLLA.

### 2.2 Setting

PRIMA will recruit inpatients from a single tertiary Vascular unit.

### 2.3 Ethical and governance approval

The full trial protocol was reviewed and approved by South East Scotland Research Ethics Committee on 03/02/2021, recognised by the United Kingdom Ethics Committee Authority (REC reference:21/SS/0013). The study is registered on ISRCTN.com (ISRCTN64207537).

### 2.4 Participants

#### 2.4.1 Eligibility criteria, informed consent, randomisation, blinding and patient withdrawal

Eligible patients will include any patient undergoing a primary MLLA, either AKA or BKA. The indication for amputation must be for end-stage chronic limb threatening ischaemia (CLTI). CLTI encompasses both patients with or without diabetes but who have peripheral arterial disease, which is severe enough to impair wound healing and thus increase the risk of MLLA. The formal definition of CLTI requires an objective evidence of PAD (cross sectional imaging +/-an ankle-brachial index (ABI)<0.4 +/-absolute Toe Pressure <30 mm Hg +/-transcutaneous arterial pressure of oxygen (TcPO2) <30 mm Hg) as well as ischaemic rest pain or tissue loss (ulceration or gangrene) either of which needs to have been present for at least two weeks.(23) Recruitment will only commence when it has been concluded from MDT discussion that amputation is the best management option by two senior vascular surgeons (consultant and/or senior vascular trainee) for the patient or the patient elects to proceed to amputation.

### 2.5 Inclusion criteria

The following patients are suitable to entry into the trial: patients aged 18 or over and undergoing a primary AKA or BKA for the symptoms resulting from CLTI (peripheral vascular disease +/-diabetes), under general anaesthesia (GA), able to consent to amputation and study participation and finally, be able to participate in assessing their pain using a Visual Analogue Scale.

### 2.6 Exclusion criteria

The following patients are not suitable for entry into the trial: patients aged below 18, undergoing amputation for malignancy or trauma, undergoing a through-knee amputation or other guillotine amputation or more proximal or revision amputations on the ipsilateral side. Patients classed as NCEPOD immediate eg; undergoing amputation for uncontrollable infection/overwhelming infection, immediately life threatening limb ischaemia are also not eligible. Patients who are unwilling/ unable to comply with the requirements for follow-up visits, who have a known allergy or contraindication to receive any constituents of the study anaesthesia, who are pregnant or lactating, or have a prior analgesia regime which includes a buprenorphine patch are also not eligible for PRIMA.

### 2.7 Informed Consent

See figure 1 which details the study schedule, outlining a timeline from enrolment to intervention and assessments in line with the SPIRIT 2013 recommendations.(22) The study flow diagram is shown in figure 2.

**Figure 1.**
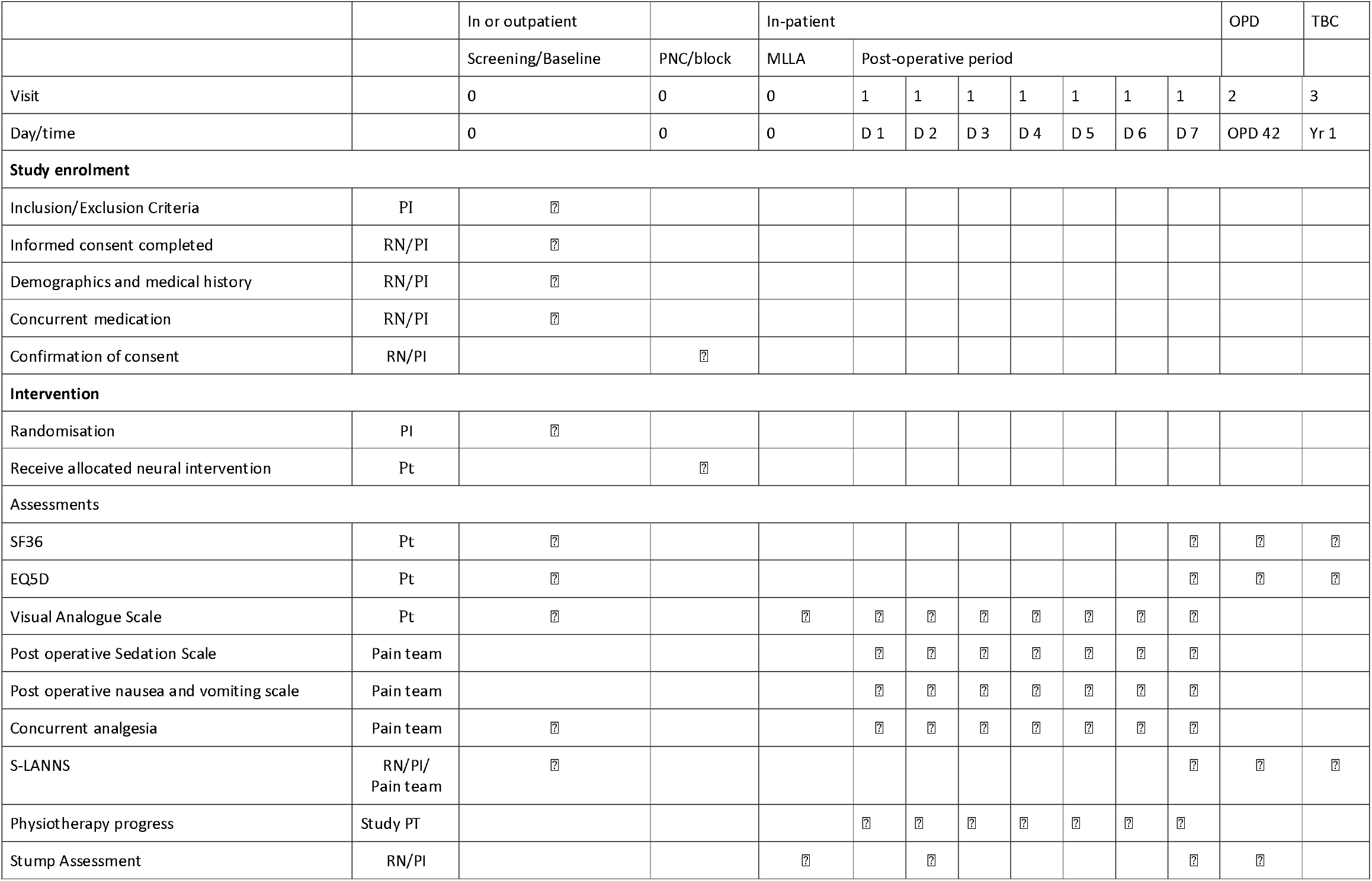
PRIMA SPIRIT schedule

**Figure 2.**
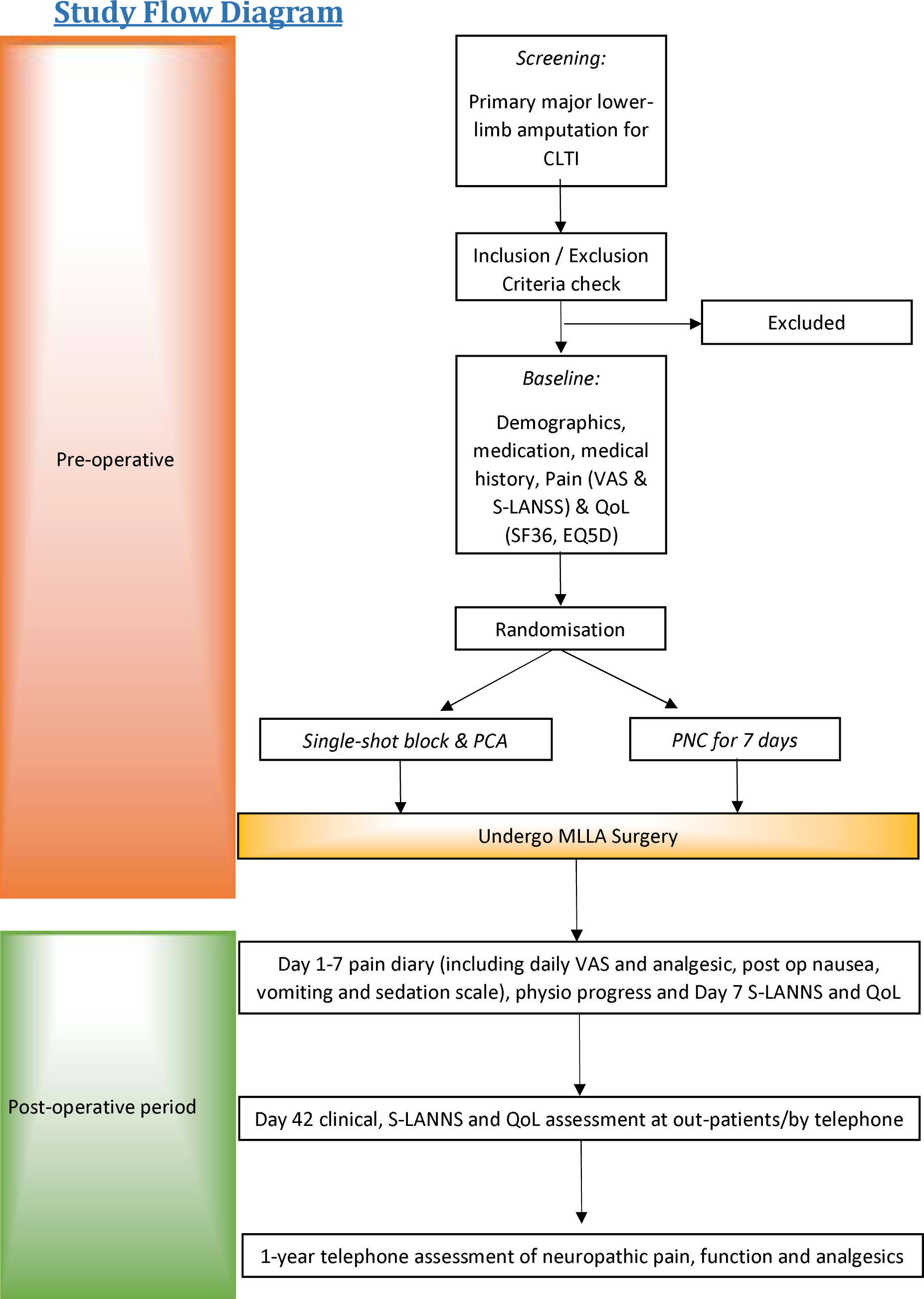
PRIMA study flow diagram

Patients planned to undergo MLLA will be screened for inclusion into the trial by members of the clinical team, against the eligibility criteria. Patients admitted via an elective or semi-elective route will be screened where possible and the study will be discussed in full at the earliest opportunity by a member of the research team. Patients will be given all the relevant information about this research study and provided with a Patient Information Sheet. Patients will be given the opportunity to think about the invitation to take part and discuss with family/ friends or other healthcare professionals if desired, prior to signing the Informed Consent Form. An anonymous log of all patients undergoing MLLA but not eligible for entry into the trial will be maintained on a trust computer, to ensure there is no selection bias.

### 2.8 Randomisation

Immediately prior to intervention, patients will be randomised to receive either a pre-incision ultrasound guided ‘single-shot’ perineural block or a PNC using a computer-generated random permuted block randomisation in a 1:1 ratio. Patients will only be randomised, if there is an anaesthetist available to who is competent to perform either intervention. A patient’s operation will not be delayed for the lack of availability of a regional anaesthetist (for example at the weekend).

### 2.9 Blinding

The nature of the intervention means double blinding is not possible. The trialists and data analysis team will be unaware of the allocation of the intervention and hence this is a single blind study.

### 2.10 Withdrawal of patients

This study will be conducted on an intention to treat basis. Patients will be withdrawn from the study at any stage if the aforementioned criteria apply or if they otherwise so desire, without prejudice to their rights to receiv appropriate treatment. PRIMA powered to allow for a 15% drop-out/ loss to follow up.

### 2.11 Audit trial conduct

The research office is subject to annual audit of one/more studies, selected at random, which could include PRIMA. Whilst this is conducted internally by the R & D quality assurance team, it is independent from study staff and sponsors.

### 2.12 Trial interventions

#### 2.12.1 Interventions, safety considerations, outcomes and timeline?

For either the intervention group or the control group, the PNCs/blocks are only sited by a core group of five Consultant anaesthetists who are established regional anaesthetists, in order to ensure standardisation. For either group, the patient has undergone a GA first. To make this study as pragmatic as possible, we have left the constituents of the GA to the discretion of the primary anaesthetist for the case. It is not possible to standardise dosing of opiates used on induction, for example, but the total amounts of intra-operative analgesics administered will be noted.

### 2.13 Above knee amputation

#### Intervention group

The participants would receive a femoral nerve catheter and a single shot sciatic nerve block.

#### Control group

The participants would receive a single shot femoral and sciatic nerve blocks and a PCA (morphine or fentanyl).

### 2.14 Below knee amputation

#### Intervention group

The participants would receive a sciatic nerve catheter and a single shot femoral nerve block.

#### Control group

The participants would receive single shot femoral and sciatic nerve blocks and PCA (morphine or fentanyl.

### 2.15 Perineural catheters

For the PNC component of the intervention group, this consists of a femoral nerve PNC for AKA or sciatic nerve PNC for BKA. The patient is supine for femoral nerve PNC or in Sim’s position for a posterior approach to sciatic nerve (Labat’s approach). The areas in question are prepped with 0.5% chlorhexidine spray and the ultrasound probe covered with a sterile covering, whilst the operator scrubs in sterile gown and gloves. The leg is then scanned and relevant nerve identified, before a small incision to the skin is made with a scalpel. A Touhy needle (Pajunk Tsui StimuLong Sono 50mm/100mm 18G Touhy needle and stimulating catheter) is inserted out of plane under ultrasound guidance, with the nerve stimulator set to 2.0mA. The correct placement is confirmed by a patella twitch for the femoral nerve and by plantar flexion of the foot/toes for the sciatic nerve. The current is reduced to threshold for loss of twitch and if 0.5mA or less, intra-neural placement is considered and the needle repositioned. Once happy with needle position, and the stimulating catheter is threaded down the needle, stimulating at 1.5mA. The catheter is threaded along the nerve until 4-6cm of catheter is next to the nerve, ensuring no loss of twitch. The catheter is loaded with up to 20 mls of 0.25% - 0.5% Levobupivacaine (max total dose per patient 2.5-3 mg/kg) and tunnelled to the anterior abdominal wall and secured to skin. A filter is applied and post-operatively the catheter is connected to a nerve catheter pump and 0.1% bupivacaine infused at 10 ml/hr for 7 days.

### 2.16 Nerve blocks

With regards to nerve blocks, which are components of both groups, for a femoral nerve block the patient is supine whilst for a sciatic nerve block the patient is in lateral or Sim’s position. The injection site is prepped with 0.5% chlorhexidine spray and the ultrasound probe covered with a sterile covering. The leg is then scanned and relevant nerve identified, before a 50mm-100mm block needle is inserted in plane under ultrasound guidance. The block is performed with up to 20 mls of 0.25% - 0.5% Levobupivacaine (max total dose per patient 2.5-3 mg/kg)

### 2.17 PCAs

The trial will utilise the standard PCA dosing in use in our trust which is morphine 1mg bolus/5 minutes with no background infusion. Fentanyl PCAs will be used in case of renal impairment or contraindication to morphine at 25microgram/5 minutes with no background infusion.

### 2.18 Pain rescue protocol

In the event that a nerve catheter is not working effectively and patients are requiring significant additional analgesia, as judged by the Acute Pain Service Specialist Nurses who will review the patients every day for 7 days or any concerned member of their clinical team, the patients will be offered a PCA. All doses of opiates utilised in the PCA will be calculated over each 24 hour period and recorded.

### 2.19 Safety considerations

Both interventions are in routine use currently, with the control group being current standard of care unless there is a regional anaesthetist available to site a PNC. Both interventions utilise levobupivacaine. A single application of levobupivacaine does not generally cause systematic side effects. The main recognised complications of a regional anaesthetic technique include local anaesthetic toxicity and nerve damage from the needle insertion. Local anaesthetic toxicity has an incidence of around 3 per 10,000 peripheral nerve blocks.(24) Hypersensitivity reactions can occur but are extremely rare. The incidence of anaphylaxis to local anaesthetics is lower than that of the commonly used skin cleaning solution chlorhexidine which has an incidence of 0.78 per 100,000 exposure, compare to no incidences of anaphylaxis to levobupivacaine recorded in the NAP 6 report.(25)

The incidence of permanent nerve damage from a regional technique is 1.5 per 10000 blocks.(26) It should however be noted that the nerves in question will be deliberately cut as part of the surgical procedure during a MLLA

As such no adverse events are expected but all adverse events will be reported.

### 2.20 Outcomes

#### Primary effectiveness outcome

The primary outcome for the study is to compare the post-procedural pain on Day 3 as evidenced utilising the patient’s visual analogue scale (VAS) score. (27, 28)

#### Secondary effectiveness outcomes

The secondary outcomes will assess other pain metrics in several ways. Firstly by documenting adjunctive pain relief medication such as opioid use, tablets, patches and neuropathic medication, preoperatively, intraoperatively and then daily for the first seven post-operative days. Secondly by daily pain scores, at baseline and then from immediately postoperatively through to day seven, using VAS. In addition, patient’s pain scores as measured using a numerical rating scale 0-10 (NRS) are routinely collected by the ward nurses every 4 hours whilst either a nerve catheter or PCA is in situ. Finally the presence of phantom limb or neuropathic pain as assessed by the S-LANNS will be captured at baseline, day 7, week 6 and 1 year.(29)

Other outcomes include the level of opioid induced sedation as measured by the Pasero Opioid-induced Sedation Scale (POSS) and the level of post-operative nausea and vomiting (PONV) as assessed by the PONV impact scale. Both of which will be asked daily for the first seven post-operative days.(30-32)

Other clinical and procedural outcomes will be documented including anaesthetic and operative times, stump healing and antibiotic use, level of engagement, ability to participate in physiotherapy and the duration of hospital stay. Finally Quality of life scores (SF36 and EQ5D) will be collected at baseline, day 7, week 6 and 1 year. (33-38)

## 3 Analysis

### 3.1.1 Sample size, statistical analysis and publication/dissemination of results

#### 3.2 Sample size

A local observational cohort study, presented at The Vascular Society of Great Britain and Ireland Annual Scientific Meeting in 2019, identified that those patients who received a PCA had a day 3 pain score, on a 0-10 numerical rating scale, of 4.8 compared to 1.2 for those who received a PNC. This is a 3.6 difference. This has been extrapolated to a 36mm difference on a VAS. From published literature, the minimum clinically important difference when using a VAS should be 13mm (95% CI 10-17mm, SD 18.3) on 100mm VAS.(39)

However the MCID is more complicated as the cohort of patients in this study will, by definition, have some degree of pain prior to their amputation (CLTI definition includes >/= 2 weeks of rest pain).(23) One review article which details mean pain scores of patients with CLTI prior to any intervention, reports scores primarily >9, with 7.9 +/-1.2 being the lowest.(40) A systematic review of empirical studies assessing the MCID in acute pain found that for each 10 mm increase in baseline pain, MCID increased by 3.1 mm (95% confidence interval, 2.8–3.5 mm, P⍰< ⍰0.001, I^2^ ⍰= ⍰0%). So for patients with high levels of initial pain (classed as >⍰170 mm) the MCIDs was 21 (20–23) mm.(41)

This being said, in order to ensure we detect a difference and allowing for changes in the magnitude, the study has been powered to detect a 25mm difference with the PNC group having a pain score of 1.5 and the single-shot group having a pain score of 3.5 with a standard deviation of 2. The sample size is 14 patients per group to see this effect size with a power of 90% and alpha of 0.05. Accommodating a 20% attrition rate the total target sample size is 34 (17 per group).

#### 3.3 Statistical analysis

This is superiority trial. We aim to establish which method of ultrasound guided pre-incision perineural analgesia is superior in terms of Day 3 patient reported pain. All data analyses will be undertaken using IBM® SPSS® Statistics.

#### 3.4 Publication and dissemination of results

PRIMA study results will be published in an international peer-reviewed journal and will be presented at a vascular national/international scientific meetings. Locally, results will presented to the regional vascular network, with a view to standardising care across the region. Social media will also be used to disseminate the results to a wider audience.

## 4 Discussion

The PRIMA study is a necessary study that addresses national patient led priorities and is supported by clinicians and allied health care professions at our centre. We have observed a positive experience of utilising pre-incision PNC. The observational cohort study mentioned as a basis to the sample size calculation, was the runner up in the BMJ 2019 awards, in the Anaesthesia and Perioperative Medicine section and was also presented at The Vascular Society of Great Britain and Ireland Annual Scientific Meeting in 2019. It included 415 patients who had undergone a MLLA between 2014 to 2018. 231 had received a PCA vs 184 who received a pre-incision US sited PNC. This identified that on average, for those patients who received a PCA, they reported a day 3 pain score on a 0-10 numerical rating scale of 4.8 compared to 1.2 for those who received a single-shot block (3.6 difference). In addition there was a 10 day difference in length of stay from 38 days for those in the PCA group to 28 days in the PNC group. This data combined with other published evidence as outlined in the background, has led to the development of PRIMA. The PRIMA study will provide level one randomised evidence to support clinical practice.

As with all studies there are some limitations. Pain assessment can be difficult and it’s noted that there are multiple methods of assessing pain, other than VAS.(28, 42, 43) VAS has been selected for use in the primary outcome measure for this study for several reasons. It’s important to note there isn’t a universally accepted pain assessment scale or tool for use in patients who have undergone amputations and none comparing the different methods, in this patient group, in the literature. The post-operative pain from an amputation is both nociceptive and neuropathic.(5) So reverting to first principals and using this knowledge, the most important reasoning behind utilising VAS is that it has been found to correlate more closely with other tools that measure both these facets of pain, than for instance the Numerical Rating Scale (NRS).(5, 44) This will be useful in our follow up, along with the use of the S-LANNS tool, to look for signs or symptoms of phantom limb pain. VAS is also commonly utilised in research and literature. A systematic review comparing studies which used VAS as well NRS and a Verbal Rating Scale(VRS), in the assessment of pain intensity in adults, showed that VAS was utilised in the largest proportion of studies (52/54) compared to NRS and VRS (32 and 39 out of 54 respectively).(43) NRS or VRS provide a discrete whole number which is not as exact as the number gained from a VAS, a continuous variable. The only other significant publication investigating ultrasound sited nerve catheter use, utilises VAS and so will be directly comparable.(18) Finally, interestingly, studies often have their results converted to mm as utilised in VAS, for the sake of comparison of results between studies.(41)

By adding S-LANSS, PONV and sedation scales as well as totalling the amounts of additional analgesia needed and physiotherapy engagement, we will provide a holistic picture of the patients in post-operative period and recovery.

The challenge with all research is to impact the lives of our patients through implementation or indeed further study. To understand this further we conducted an international survey ‘*PReliMinAry*’ in 2021 to identify any potential barriers for units utilising pre-incision US guided PNC. One barrier cited was ‘the lack of expertise to do so’. (4) Of the 60 units who responded to questions surrounding their analgesic regimes in MLLA, only 77% (n=46) were currently able to offer pre-incision US guided PNC as an option, versus 90% (n=54) for surgical sited PNC. 62 of 76 survey respondents (86%) said they would be happy to randomise patients in a trial to pre-incision US guided PNC but the commonest reason for those who selected maybe or no was that there is a lack of expertise to actually perform the technique. That training can be difficult to access and justify without good quality evidence behind a technique, which is another reason behind PRIMA.

Whilst the number to recruit is only small number, there would be a potential to springboard to a larger RCT.

## Data Availability

Data is yet to be published but will be in due course.

## 5 Acknowledgements

PRIMA has been supported from the start by a broad multi-disciplinary team. We acknowledge and thank the contributions to date from the following:

Vascular research nurses: Leslie Bremner, Jenny Fairbairn and Noala Parr

Data manager: Anthony Robinson

Pain team nurses: Katherine Ashton, Jenny Houston and Angela Telford Consultant anaesthetists: Rachael Bird, Martin Jones, Rita Singh and Iain Walker

Physiotherapists: Katie Bell, Tamsin Jeffery, Alexander Kermode and Catherine McGarry

## Notes

Funding: This work was supported by a pump priming grant from the Royal College of Surgeons of Edinburgh (RCSEd), grant number N/A. The RCSEd had no role in the study design, writing of this report of decision to submit this article for publication.

### Competing Interest Statement

The authors have declared no competing interest.

### Clinical Trial

ISRCTN64207537

### Author Declarations

South East Scotland Research Ethics Committee gave ethical approval for this work on 03/02/2021. REC reference:21/SS/0013.

